# Multi-polygenic scores in psychiatry: from disorder-specific to transdiagnostic perspectives

**DOI:** 10.1101/2022.05.30.22275563

**Authors:** Yingjie Shi, Emma Sprooten, Peter Mulders, Janna Vrijsen, Janita Bralten, Ditte Demontis, Anders D. Børglum, G. Bragi Walters, Kari Stefansson, Philip van Eijndhoven, Indira Tendolkar, Barbara Franke, Nina Roth Mota

## Abstract

The dense co-occurrence of psychiatric disorders questions the categorical classification tradition and motivates efforts to establish dimensional constructs with neurobiological foundations that transcend diagnostic boundaries. In this study, we examined the genetic liability for eight major psychiatric disorder phenotypes under both a disorder-specific and a transdiagnostic framework. In a deeply-phenotyped sample (n=513) consisting of 452 patients from tertiary care with mood disorders, anxiety disorders, attention-deficit/hyperactivity disorder (ADHD), autism spectrum disorders (ASD), and/or substance use disorders (SUD) and 61 unaffected comparison individuals, we derived subject-specific multi-base polygenic risk score (PRS) profiles and assessed their associations with psychiatric diagnoses, comorbidity status, as well as cross-disorder behavioral dimensions. High PRS for depression was unselectively associated with the diagnosis of SUD, ADHD, anxiety disorders, mood disorders, and the comorbidities among them. In the dimensional approach, four distinct functional domains were uncovered, namely the negative valence, social, cognitive, and regulatory systems, closely matching the major functional domains proposed by the Research Domain Criteria (RDoC) framework. Critically, the genetic predisposition for depression was selectively reflected in the functional aspect of negative valence systems but not others. This study highlights a misalignment between current psychiatric nosology and the underlying psychiatric genetic etiology, and underscores the effectiveness of the dimensional approach in both the functional characterization of psychiatric patients and the delineation of the genetic liability for psychiatric disorders.

## Introduction

Psychiatric disorders are among the most common, disabling, and costly diseases in humans ^[1]^, and yet, science falls short in understanding their etiopathogenesis. Conventional diagnostic frameworks, represented by the Diagnostic and Statistical Manual of Mental Disorders (DSM) ^[2]^ and International Classification of Diseases (ICD) ^[3(p. 1)]^, have often been employed as the scaffolding for mechanistic investigation and risk factor identification within case-control designs. However, limitations of adopting such discrete diagnostic frameworks in the research context have been well-recognized, and distinct boundaries among diagnostic categories are challenged by the misalignment with patient profiles. Specifically, substantial differences in symptom profiles ^[4]^ (i.e., phenotypic heterogeneity) as well as neuronal features ^[5,6]^ (i.e., biological heterogeneity) exist within the same diagnostic category, while patients with differently classified disorders could converge on overlapping symptomatology and/or pathological pathways ^[7]^. The frequent observation of co-occurrence of multiple psychiatric disorders in clinical practice is closely tied with such heterogeneity and overlap. The high prevalence of psychiatric comorbidity ^[8]^ and the associated poorer clinical outcome requires research to move beyond a single diagnosis and focus on the identification of transdiagnostic mechanisms. Several initiatives proposing dimensional alternatives have been established, such as the NIMH Research Domain Criteria (RDoC) ^[9]^ and the Hierarchical Taxonomy of Psychopathology (HiTOP) ^[10]^. In particular, the RDoC framework aims to explicate the neurobiological foundation of psychopathology using transdiagnostic bio-behavioral domains, namely Negative Valence Systems, Positive Valence Systems, Cognitive Systems, Systems for Social Processes, Arousal/Regulatory Systems, and Sensorimotor Systems ^[11]^.

Recent psychiatric genetic studies have confirmed the overlapping genetic architecture among different disorders, pointing towards shared genetic substrates. The team efforts coordinating large-scale genome-wide association study (GWAS) meta-analyses have identified common genetic variations contributing to psychiatric disorders such as major depressive disorder (MDD) ^[12]^, anxiety disorders (ANX) ^[13]^, attention-deficit/hyperactivity disorder (ADHD) ^[14]^, autism spectrum disorder (ASD) ^[15]^, bipolar disorder (BP) ^[16]^, and schizophrenia (SCZ) ^[17]^. Building upon the GWAS knowledge base, genetic sharing among psychiatric disorders has been evaluated, which revealed substantial genetic overlap at the genomic level ^[18]^ from which over a hundred genetic variants exerting pleiotropic effects on more than one disorders could be identified ^[19,20]^. The identification of the polygenic architecture and effect sizes carried by individual single nucleotide polymorphisms (SNPs) enables researchers to quantify the combined genetic susceptibility to disorders in the form of polygenic risk scores (PRSs), whose usefulness has been shown in risk prediction for common diseases ^[21]^ and treatment outcome prediction ^[22]^, in identifying cross-disorder associations ^[23]^, but also in investigating complex traits that are relevant to multiple disorders ^[24]^. However, the relations of PRS for different psychiatric disorders with transdiagnostic traits have rarely been investigated in clinically rather typical, highly comorbid cohorts.

In the current study, we applied polygenic score analysis in a naturalistically recruited psychiatric cohort with high clinical complexity and comorbidity with two objectives. Under the conventional DSM-based framework, we aimed to assess the validity and specificity of PRS of major psychiatric disorders for different diagnostic outcomes. The PRSs were derived from the most recent and well-powered GWASs on MDD ^[12]^, ANX ^[13]^, ADHD ^[14]^, ASD ^[15]^, BP ^[16]^, SCZ ^[17]^, depression (DEP) ^[25]^, and cross-disorder diagnoses (Cross-disorder) ^[20]^ for patients from the recruited MIND-SET cohort and individuals free of any psychiatric disorder. Under a transdiagnostic dimensional framework, we explored the polygenic risk mapping to symptom and trait dimensions. For the latter, we first performed an exploratory factor analysis to explore latent structures in a broad range of psychopathological assessments of psychiatric, personality, and psychological symptoms and traits. Individuals’ representations on the derived functional dimensions were then examined with regard to the multi-PRS and their comorbidity status.

## Methods

### The MIND-SET cohort

The study sample MIND-SET cohort ^[26]^ was first established in Nijmegen, The Netherlands, in 2015. Recruited from the outpatient population of the department of Psychiatry at Radboud University Medical Center, the sample is composed of adult (≥ 18 years) patients with a clinical diagnosis in at least one of five disorder categories (i.e., mood disorders, anxiety disorders, ADHD, ASD and/or substance-related disorders). Individuals with current psychosis, IQ lower than 70, or inadequate command of the Dutch language were excluded from the study. A comparison group with similar demographics as the patients but free of any previous or current psychiatric disorders were recruited from the local population. Written informed consent was obtained from all participants included in the study. The study was approved by the local medical ethics committee (Commissie Mensgebonden Onderzoek Arnhem-Nijmegen).

### Disorder diagnosis

The diagnosis of patients was confirmed by a trained clinician during a structured interview. The absence of a lifetime psychiatric diagnosis in the control group was confirmed using the same diagnostic instruments via telephone interview. Mood disorders and anxiety disorders were diagnosed by means of the Structured Clinical Interview for DSM-IV Axis I Disorders (SCID-I) ^[27]^. For ASD and ADHD, a diagnosis was provided based on the results from the Dutch Interview for the Diagnosis of ASD in adults (NIDA) ^[28]^ and Diagnostic Interview for ADHD in adults (DIVA) ^[29]^, respectively. Substance use disorders (SUD) were diagnosed according to DSM-5 criteria and an adapted version of the Measurements in the Addictions for Triage and Evaluation (MATE) ^[30]^. Patients with schizophrenia and other psychotic disorders based on SCID-I were excluded. A detailed overview of the individual diagnoses included in each abovementioned disorder category is presented in **Supplementary Table S1**. Individuals were identified as having comorbid disorders if they had diagnoses that fell into more than one of the disorder categories of mood disorders, anxiety disorders, ADHD, ASD, and SUD (i.e., only comorbidity between disorder categories was considered).

### Symptoms/trait questionnaires and exploratory factor analysis

A rich test battery was utilized to characterize the study sample with regard to disorder-related psychiatric symptoms, personality traits, and other psychological traits. **Supplementary Table S2** provides an overview of these questionnaires and their subscales used in the subsequent factor analysis. Further, the level of dysfunction in daily life of participants was assessed. The self-report World Health Organization Disability Assessment Schedule (WHODAS) 2.0 ^[31]^ was used to measure disability in six domains of functioning (i.e., cognition, mobility, self-care, getting along, life activities, and participation), and the Outcome questionnaire-45 (OQ-45) ^[32]^ was used to measure subjective experiences and social functioning in domains of symptom distress, interpersonal relations, and social role.

To derive transdiagnostic dimensions measured by the scales in **Supplementary Table S2**, exploratory factor analysis (maximum likelihood estimation, oblique rotation) was performed based on 387 participants (327 patients) who completed the entire test battery. The same analysis was previously conducted in a subset of participants from the same cohort, as described in ^[33]^. A four factor solution outperformed the simulated eigenvectors in the parallel analysis (**Supplementary Figure S1)** ^[34]^, which was in line with the scree-plot.

### Base GWAS datasets

We used the most recent and well-powered GWASs for both single psychiatric categories, including MDD ^[12]^, ANX ^[13]^, ADHD ^[14]^, ASD ^[15]^, BP ^[16]^, and SCZ ^[17]^, as well as broader disorder phenotypes, including DEP ^[25]^ and Cross-disorder ^[20]^ as the bases to derive PRSs for each participant of the MIND-SET cohort (see **Supplementary Table S3** for an overview of the datasets). The DEP GWAS included both cases who had clinically ascertained diagnosis of major depressive disorder (43k, as described in ^[12]^) and cases of ‘broad depression’ who reported help-seeking behavior for mental health difficulties (128k, as described in ^[35]^). All summary statistics from these datasets, except for ADHD, are publicly available. Only biallelic SNPs with minor allele frequency (MAF) higher than 0.01 and INFO score higher than 0.9 (if available) were retained for subsequent analyses.

### Genotyping, quality control, and imputation of the target dataset

The MIND-SET cohort was genotyped on the Infinium Global Screening Array (GSA-24 v3.0). The bioinformatics pipeline Ricopili ^[36]^ (version from 2019_Oct_15.001), developed by the Psychiatric Genomics Consortium (PGC) Statistical Analysis Group, was employed to perform quality control and imputation on the genotyped data. To comply with the informed consent of the participants of the MIND-SET study, we removed variants known to be causative of diseases or disorders (i.e., pathogenic and likely pathogenic) in the genotyped data. We first excluded the variants within the pathogenic genes from the most recent list of ACMG (SF v2.0) genes recommended for return of secondary findings in clinical sequencing ^[37]^. This step was conducted before performing any (pre-)processing on the genotyped sample in order to eliminate their impact on the imputation of other variants. MAF filter of 0.01 was applied after imputation to further remove the rare pathogenic variants that were imputed back to the data.

Several filters were applied to exclude individuals and SNPs of low genotyping quality: SNP call rate < 0.95 (pre-filter), subject call rate < 0.98 for both cases and controls, autosomal heterozygosity deviation (F_HET_) outside ± 0.20, sex mismatch between genetic and phenotypic data, SNP call rate < 0.98, differences in SNP missingness between cases and controls > 0.02, SNP Hardy-Weinberg equilibrium (HWE) p < 10^−6^, and invariant SNPs. To address population stratification, we performed principal component analysis (PCA) on the preprocessed data and removed the genetic outliers that were more than three standard deviations beyond the center of the European reference cluster in the 1000 Genome Project ^[38]^. Further, overlapped/related individuals with pi-hat values greater than 0.2 were removed.

The imputation process was implemented by combining the Ricopili structure with the Michigan Imputation Server ^[39]^ (https://imputationserver.sph.umich.edu). After alignment with the reference panel, the genotypes from 22 autosomal chromosomes were phased (Eagle v2.4) and then imputed (Minimac 4) on the online server. We used the largest reference panel available, the Haplotype Reference Consortium (HRC) panel (r1.1 2016, http://www.haplotype-reference-consortium.org/), which consists of 39 million SNPs from 32,470 samples of predominantly European ancestry. The imputed data was then integrated back to the Ricopili structure and best guess genotypes were generated. PCA was performed again on the best guess genotypes, from which the first four derived principal components (PCs) were included as covariates in the subsequent polygenic score analyses, in addition to age and sex.

### Polygenic risk score calculation and association test

For each individual in the MIND-SET cohort, PRSs for the eight GWAS bases mentioned in the above section were created using PRSice 2.3.3 ^[40]^. Clumping was performed using a linkage disequilibrium r^2^ threshold of 0.1 and a sliding window of 250 kb to ensure independence among SNPs. *A priori* sets of ten P-value thresholds (P_T_; i.e., 5e-8, 1e-6, 1e-4, 0.001, 0.01, 0.05, 0.1, 0.2, 0.5, 1) were applied to the base GWASs to compute different genome-wide PRSs for each subject, and the best-fit PRS P_T_ (i.e. the most strongly associated PRS P_T_) for each outcome of interest (i.e., diagnostic outcomes, factor dimensions) was identified and retained. To avoid overfitting, we computed empirical p-values by performing 10,000 permutations ^[40]^. Additional Bonferroni correction (α = 0.05/(8*5) = 0.00125) was applied to account for the multiple tests with different base disorders/traits and the outcome disorder statuses.

PRSs were used as predictors in both simple and multiple logistic regression models for diagnostic outcomes, and simple linear regression for factors scores. The proportion of variance explained by the PRSs in all outcomes was estimated by Nagelkerke’s pseudo-R^2^, computed as the difference between the R^2^ of the full model, containing the PRS and the covariates (i.e., age, sex, and the first four PCs), and the R^2^ of the null model, containing only the covariates. One-way ANOVAs were applied to test the differences in PRSs and factor loadings among groups with different comorbidity statuses.

## Results

### Diagnoses and comorbidities in the MIND-SET cohort

A total of 513 individuals of European ancestry were included in the study: 452 had at least one diagnosis of mood disorder, anxiety disorder, ADHD, ASD, and/or SUD, and 61 were sex- and age-matched unaffected individuals. An overview of the refined diagnoses within each disorder category is presented in **Supplementary Table S1**. Among the patients, 80% (n=360) had at least one diagnosis in the mood disorders spectrum, 33% (n=147) had at least one anxiety disorder, 38% (n=171) had ADHD, 27% (n=121) had ASD, and 27% (n=121) had SUD. Psychiatric comorbidity was highly prevalent in the MIND-SET cohort: 70% of the patients fell into at least two diagnostic categories, and 28% into three or more. As shown in **Figure 1A**, mood disorders in combination with anxiety disorders, ADHD, or SUD were among the most common comorbidities in the current cohort.

**Figure 1.**
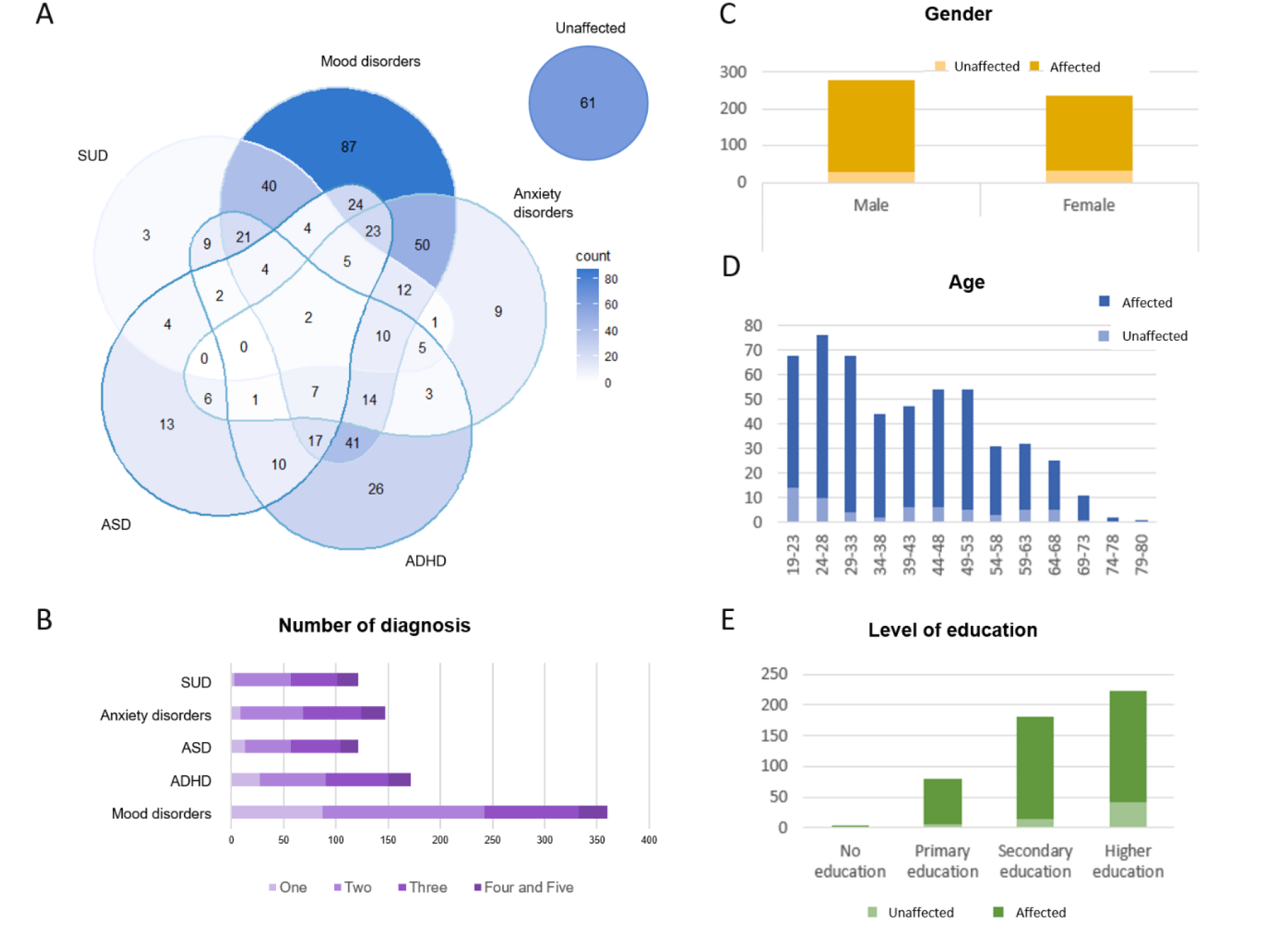
MIND-Set participant characteristics, psychiatric diagnoses, and comorbidity status. Venn diagram of the five disorder groups (**A**) and the summary of the number of groups patients fell into (**B**). Multiple diagnoses within the same disorder group were regarded as one. The distribution of gender (**C**), age (**D**), and level of education (**E**) of cases and the unaffected comparison group. There were no significant gender or age differences (t = 1.04, p = 0.297; t = −0.39, p = 0.696) between the two groups; the affected group had lower levels of education than the unaffected comparison group (Chi-squared = 15.32, p = 0.002).

### Multi-base PRS and disorder diagnostic status

PRSs computed based on the broadly-defined phenotype of depression (i.e., DEP-PRS) were significantly associated not only with mood disorder status, but - even to a larger extent - with SUD, ADHD, and anxiety disorders (**Table 1**, all at P_T_ = 0.01). By contrast, neither single disorder-based PRSs (incl. MDD, ANX, ADHD, ASD, BP, SCZ) nor the cross-disorder PRSs significantly explained the diagnostic status of any disorder category after Bonferroni correction. The PRS distributions of individuals within each disorder category and unaffected comparisons are depicted in **Figure 2A** for DEP and in **Supplementary Figure S2** for other PRSs. Combining the genetic risks across different base disorders, we present in **Figure 2B** the multi-axis genetic risk profiles for the affected and unaffected groups. Since group differences in several disorder categories were found also for the ADHD-, ANX-, and SCZ-PRSs at an uncorrected significance threshold (**Table 1**), we tested whether adding these multi-disorder PRSs to the model would improve prediction for disorder outcome compared to the DEP-PRS alone. Using all eight PRSs as predictors, the multiple regression model explained 3.07% - 7.47% more variance than the model with DEP-PRS as the single predictor (**Figure 2C**), but did not yield statistically significant improvement on the model fit for the disorder outcomes (**Supplementary Table S4**).

**Table 1.** Variance explained (pseudo-R^2^ [p values]) in diagnosis status using PRS for different disorders. The proportion of variance explained by each PRS in each of five psychiatric disorder diagnoses was estimated by Nagelkerke’s pseudo-R^2^, computed as the difference between the R^2^ of the single PRS model, containing one PRS and the covariates (i.e., age, sex, and four PCs), and the R^2^ of the null model, containing only the covariates.

| Phenotype | MDD-PRS | ANX-PRS | ADHD-PRS | ASD-PRS | BP-PRS | SCZ-PRS | DEP-PRS | Cross-disorder-PRS |
| --- | --- | --- | --- | --- | --- | --- | --- | --- |
| **Mood disorders** | 0.0198<br>[0.153] | *0.0320*<br>[0.041] | 0.0156<br>[0.221] | 0.0097<br>[0.547] | 0.0216<br>[0.100] | 0.0218<br>[0.086] | **0.0826**<br>[0.0002] | 0.0232<br>[0.090] |
| **Anxiety disorders** | 0.0407<br>[0.084] | 0.0336<br>[0.146] | 0.0166<br>[0.418] | 0.0031<br>[0.985] | 0.0387<br>[0.080] | *0.0507*<br>[0.027] | **0.1253**<br>[0.0003] | 0.0447<br>[0.062] |
| **ADHD** | 0.0273<br>[0.171] | *0.0471*<br>[0.038] | *0.0577*<br>[0.016] | 0.0074<br>[0.841] | 0.0321<br>[0.106] | 0.0301<br>[0.104] | **0.1271**<br>[0.0003] | 0.0279<br>[0.157] |
| **ASD** | 0.0250<br>[0.270] | 0.0357<br>[0.150] | 0.0404<br>[0.093] | 0.0032<br>[0.984] | 0.0217<br>[0.293] | 0.0427<br>[0.062] | *0.0597*<br>[0.023] | 0.0286<br>[0.200] |
| **SUD** | 0.0195<br>[0.407] | *0.0570*<br>[0.039] | 0.0304<br>[0.181] | 0.0252<br>[0.331] | 0.0295<br>[0.178] | 0.0197<br>[0.296] | **0.1338**<br>[0.0007] | 0.0318<br>[0.176] |

**Note.** Associations that exceeded Bonferroni-corrected threshold of $p < 0.05/(5 \text{ phenotypes} * 8 \text{ base disorders}) = 0.00125$ are labeled in **bold**, and those exceeding the uncorrected threshold of $p < 0.05$ are labeled in *italic*. MDD - major depressive disorder; ANX - anxiety disorders; ADHD - attention-deficit/hyperactivity disorder; ASD - autism spectrum disorders; BP – bipolar disorder; SCZ – schizophrenia; DEP – depression; SUD – substance use disorders.

**Figure 2.**
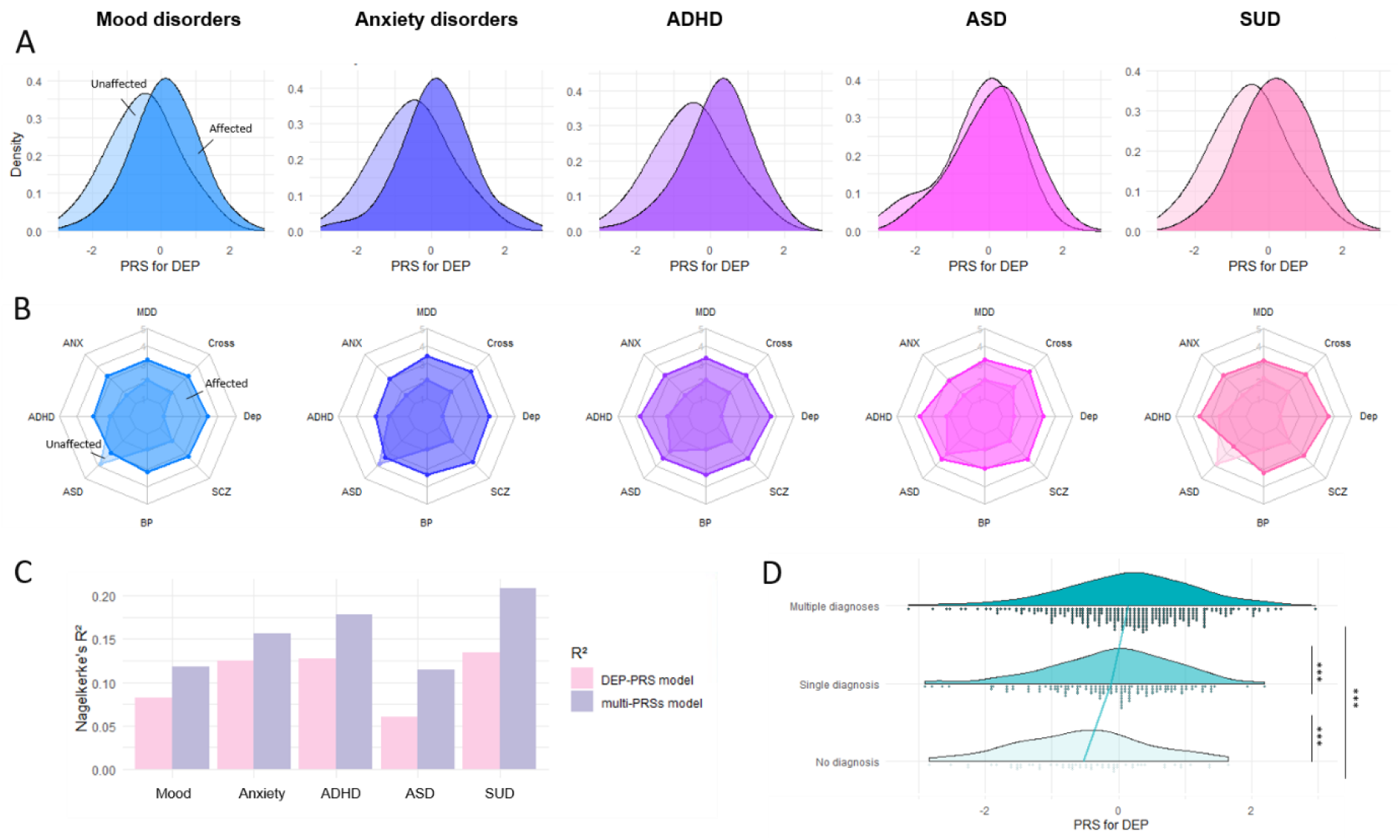
Multi-base PRS profile for different disorder categories and comorbidities. **A**. PRS distributions for broad depression of unaffected (light color) and patient samples (dark color) of the disorder shown under each subplot. See **Figure S2** for PRS distributions of MDD, ANX, ADHD, ASD, BP, SCZ, and Cross-disorder. **B**. Eight-axis PRS profiles for each disorder group, with each axis representing the PRS based on one GWA study. PRSs were constructed using the P_T_ that yielded the strongest associations with the outcome of interest. **C**. Variance explained by DEP-PRS as the single predictor in comparison to eight PRS predictors. R^2^ for the null model (age, sex, and four PCs) has been subtracted from both model. **D**. Group differences in PRS for depression with regard to individuals’ number of diagnoses (group with no diagnosis N = 61, single diagnosis N = 138, multiple diagnoses N = 314). The reference line connects the average value for each group. *** p < 0.001.

To further test whether DEP-PRS was related to disorder comorbidity status, we compared the DEP-PRS among groups of unaffected individuals, patients with a single disorder and the comorbid group (**Figure 2D**). The results suggested a significant overall DEP-PRS effect on comorbidity status (F = 12.95, *p* = 3.3e-06): patients with comorbid conditions had higher DEP-PRS compared to patients with only one diagnosis (t = 2.50, *p* = .033), which in turn had higher DEP-PRS than unaffected individuals (t = 2.78, *p* = .015).

### Data-driven functional dimensions

Converging a wide array of psychopathology assessments into cross-disorder constructs, the factor analysis of 31 (sub)scales of psychiatric, personality, and psychological traits measured in MIND-SET (**Supplementary Table S2**) yielded four factors, which together explained 67.3% of the variance (KMO = 0.948, Bartlett’s test p < .001). These four factors matched the previous finding using a subsample of the same cohort with a highly similar component matrix ^[33]^, in which the interpretation of the factors roughly corresponded to previously defined RDoC domains ^[9,11]^ (**Figure 3A**): the first factor related to negative thinking, emotions, and poor self-concept across instruments (i.e., RDoC negative valence systems); the second factor summarized difficulties in social functioning (i.e., RDoC social processes); the third factor described cognitive abilities (i.e., RDoC cognitive systems); the last factor related to the ability in regulation and inhibition (i.e., RDoC arousal/regulatory systems). To evaluate the relevance of the derived factors to individuals’ functioning and disabilities, we assessed their relationship with self-rated quality of life measured using OQ-45 and WHODAS 2.0 scales. We found that all four factors had significant positive regression weights for the outcome of individuals’ subjective distress and social dysfunction measured in OQ-45, and the first three factors had significant positive regression weights for the outcome of overall disability measured with WHODAS 2.0 (**Supplementary Table S5**); together, they explained the outcomes with an adjusted R-squared of 0.81 and 0.65, respectively. Different disorder categories were represented by distinct factor profiles which resembled their clinical presentations (**Supplementary Figure S3**). For example, patients with ADHD loaded highly on the dysfunction in cognitive and arousal/regulatory systems, whereas patients with ASD had higher dysfunction loading in the social processes. Compared to unaffected individuals, all patients scored higher on the loading of dysfunction on all factors (**Figure 4, Supplementary Table S6**). Compared to the group with only one diagnosis, the comorbid group had significantly higher loadings for dysfunction after multiple testing correction (α_adj_ = .05/4 = .0125) for negative valence systems (t = 4.93, *p* = 3.56e-06), social processes (t = 4.03, *p* = .002), and arousal/regulatory systems (t = 4.32, *p* = 5.6e-05). For cognitive systems, there was no significant difference in loading between groups with single and multiple diagnoses (t = 0.48, *p* = 0.878).

**Figure 3.**
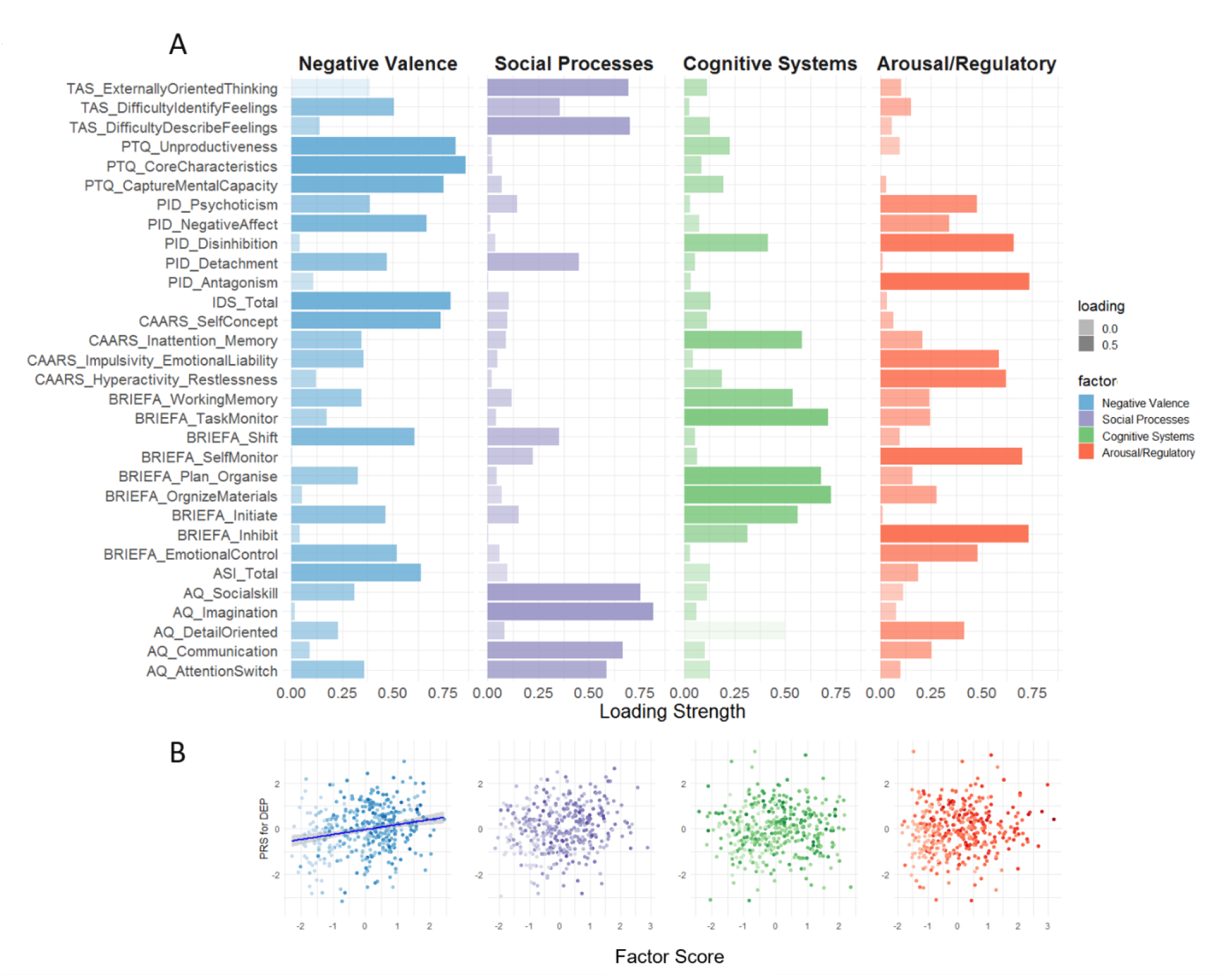
Rotated component matrix of four factors resulting from factor analysis of psychopathology measurements. **A**. Four factors were retained after parallel analysis and were interpreted in the column headers. The Y-axis shows the (sub)scales included in the analysis following the naming scheme – ‘questionnaire name abbreviation_subscale’. Please consult **Table S2** for a detailed list of the questionnaires included in the EFA. The component matrix contains the factor loadings (Pearson correlations between items and components) on each subscale with color intensity corresponding to the loading strength. **B**. Individuals’ PRS for depression in relation to their scoring on each factor dimension. Line of best fit is plotted for the negative valence factor, which is significantly correlated with PRS for depression. Color intensity is scaled according to the number of diagnoses.

**Figure 4.**
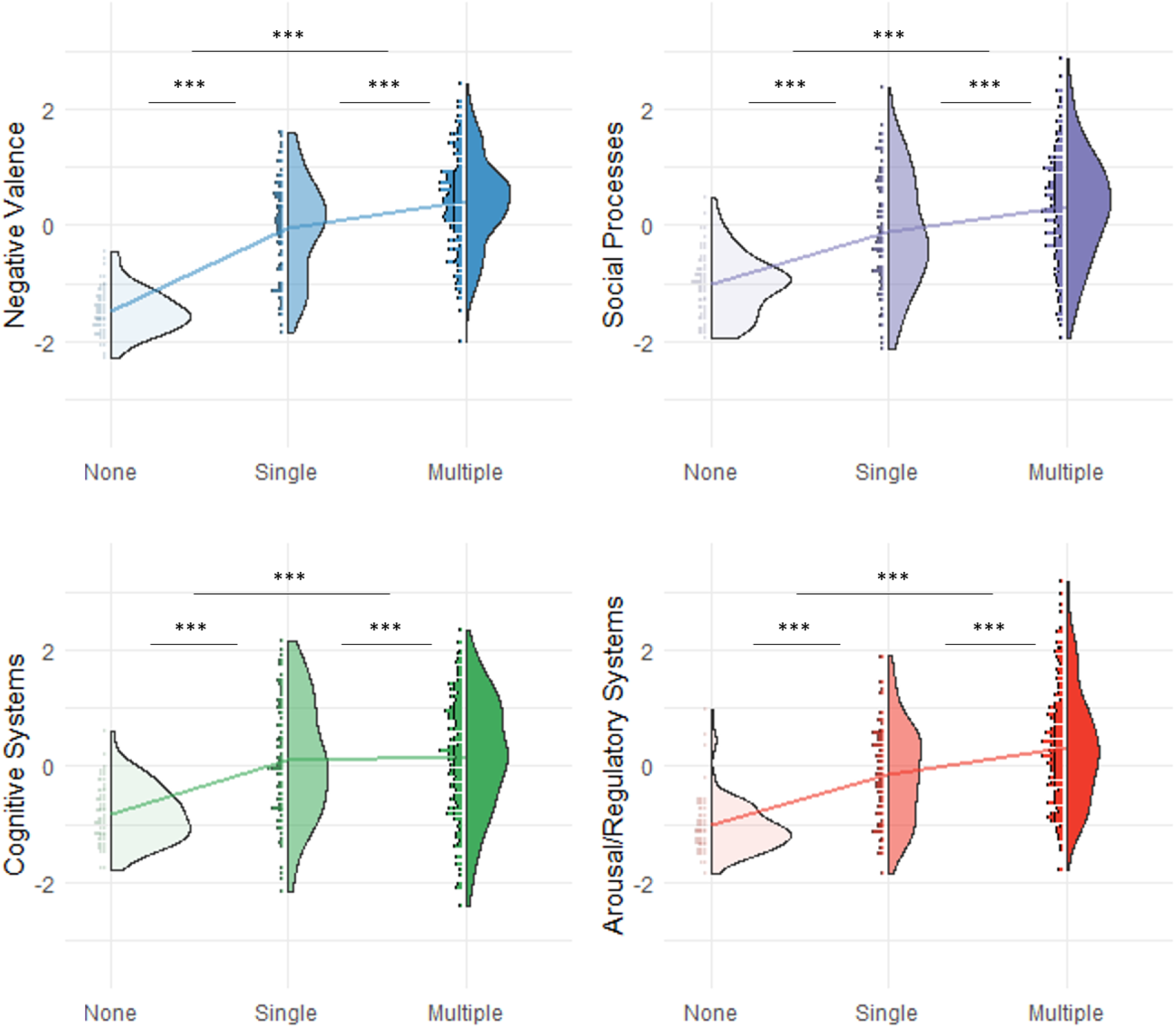
Group differences in factor loading with regard to comorbidity status. Individuals’ loadings for four factor dimensions were stratified by whether the individual had no disorder diagnosis (N = 60), single diagnosis (N = 95), or more than one diagnoses (multiple; N = 232). The line in each subplot connects the mean of the three groups. Higher score indicates higher dysfunction. The post-hoc tests were performed using Tukey method. *** *p* < .001; n.s. not significant.

Given the broad relevance of DEP-PRS for mood and neurodevelopmental disorders, we further tested whether this PRS was associated with specific aspect(s) of behavioral functioning. We found that the DEP-PRS was significantly associated with the negative valence dimension (**Figure 3B**, R^2^ = 0.045, *p*_*emp*_ = .006 at P_T_ = 0.01), but not with social processes (R^2^ = 0.020, *p*_*emp*_ = .022 at P_T_ = 0.2), cognitive systems (R^2^ = 0.001, *p*_*emp*_ = .948 at P_T_ = 0.001), or arousal and regulatory systems (R^2^ = 0.012, *p*_*emp*_ = .132 at P_T_ = 0.001). To confirm that this association was not driven by the deviation between the case and control distributions, we excluded the comparison group and found that this association still existed when including only affected individuals (R^2^ = 0.022, *p*_*emp*_ = .036 < .050).

## Discussion

Bringing genetic metrics derived from case-control samples into a highly comorbid clinical cohort, our study provided a real-world assessment of the validity and specificity of psychiatric polygenic risk scores, with regard to both disorder-specific and transdiagnostic outcomes. Multi-base PRS analysis revealed that the DEP-PRS outperformed all other PRSs and can significantly predict the diagnostic statuses of SUD, ADHD, anxiety disorders, and mood disorders. We reproduced four transdiagnostic dimensions derived from a diversity of psychology and psychopathology measurements and revealed that one specific dimension - the negative valence system - was selectively associated with DEP-PRS.

Our association analyses with DSM diagnoses showed that the genetic propensity for a broadly-defined depression phenotype (i.e., DEP) can significantly predict disorder statuses outside of the mood disorders spectrum. With more than 75% of cases identified based on ‘minimal phenotyping’ (i.e., a positive answer to the question “Have you seen a GP/psychiatrist for nerves, anxiety, stress or depression?”), the original GWAS on which the DEP-PRS was computed (excluding 23andMe cohort) ^[25]^ is phenotypically much broader than GWAS using clinically ascertained (major) depression ^[12]^ and statistically more powerful given its sample size. Previous studies have addressed the fact that such broad phenotyping approaches might identify a genetic architecture that is not specific to the clinical form of MDD ^[41]^, but noticed that those can be highly useful for risk prediction and risk factor identification - especially given the convenience to reach large sample sizes ^[42]^.

Consistent with the previous factor analysis results from a smaller overlapping sample ^[33]^, we observed four transdiagnostic domains (i.e., negative valence, social, and arousal/regulatory systems) among a broad battery of psychiatric, personality, and psychological assessments, which together explained up to 81% of individuals’ subjective experience of their functioning and disability. Importantly, the finding that DEP-PRS was selectively linked to the functioning of the negative valence system suggests that a shared domain of functioning could underlie its associations with multiple disorders. This underscores the importance of a paradigm shift in mechanistic investigations towards dimensional constructs that are data-driven and acknowledge the intertwined nature of different categorized psychiatric disorders. Uncovering the latent dimensions of psychopathology from dense symptom- and trait-level data, on the one hand, will help to identify individuals’ unique (dys-)functional profile and enable targeted interventions for specific functional spectra; on the other hand, they can provide neurobiological studies with an improved scaffolding to investigate underlying pathogenic processes. Although attempts of genomic enquiry on dimensional traits established in a data-driven fashion are still scarce, efforts of GWAS on theory-based psychopathology traits (e.g., extracted from clinical notes ^[43]^, neurocognitive tests ^[44]^, self-report assessments ^[45]^) are blooming and provide important leads for follow-up causal assessments.

With its broad relevance, the DEP-PRS explained different diagnostic outcomes in our cohort to a higher magnitude than previously found in other cohorts ^[25]^. While an independent sample is needed to further validate the predictive accuracy, we postulate that the contrast between highly severe and comorbid cases recruited from a specialized academic hospital and a ‘clean’ comparison group free of any psychiatric history could amplify the predictive ability of PRS. This could lead to a larger R^2^ compared to previous reports where patients may have had less complex, less comorbid clinical profiles, and controls were not always screened for other major psychiatric disorders ^[25]^. Furthermore, we recognize that the composition of disorders of the current cohort might align well with the constellation of psychiatric characteristics probed by such ‘minimal phenotyping’ ^[41]^ definition of the broad depression PRS, which will also give rise to a more effective PRS.

The heterogeneity in the current study sample has been maximized to span a large spectrum of mental health and functioning, ranging from individuals free of mental health complaints to tertiary care patients with multiple diagnoses. Unlike previous studies where comorbid conditions were either ignored (i.e., participants are unscreened for other disorders) or treated as confounding variables or exclusion criteria, we addressed the topic of psychiatric comorbidity explicitly and characterized it on both the genetic and cross-disorder behavioral scales. We showed that individuals displaying comorbidity were bearing higher genetic liability and displayed a higher degree of dysfunction in most functional aspects. This adds functional and biological evidence to the large body of comorbidity literature showing phenotypic associations between psychiatric comorbidity and higher severity and more chronicity of impairment (e.g., ^[46,47]^). Rather than regarding psychiatric disorders as distinct entities that deserve separate treatments on top of each other, it is crucial to acknowledge the shared underlying vulnerability factors and etiopathogenesis that push individuals to the higher end of the psychopathology spectrum.

The present study provides a thorough assessment of the validity and specificity of PRSs for major psychiatric disorders against categorical as well as dimensional outcomes by exploiting a clinically well-assessed cohort spanning a wide spectrum of psychopathology. However, several limitations need to be taken into account when interpreting the results. First, while a permutation procedure was performed to adjust association p-values, the observed phenotypic variance explained (R^2^) requires an independent sample to evaluate the predictive accuracy of the PRSs. Second, our patient sample was enriched with mood disorder either as a single diagnosis or as a comorbidity, which could limit the generalizability of our results for the other, non-mood related disorders to samples focused on those other disorders as a standalone diagnosis. Third, only comorbidities among the defined non-psychotic disorder categories (as opposed to within disorder categories) were considered, which may yield an incomplete picture of the functional and genetic characterization of psychiatric comorbidity.

In conclusion, our polygenic scoring analysis revealed low specificity to psychiatric disorders as defined by conventional classification systems, but enhanced specificity to data-driven functional domains. Domain-based genetic analyses targeting traits and symptoms not restricted to a single disorder or clinically ascertained group could help reduce the clinical and biological heterogeneity of the study sample and enable more fine-grained mapping to the biological basis of psychopathology at different levels. It also supports further initiatives of targeted treatments based on neurocognitive domains that eventually can provide an important avenue for psychiatric interventions.

## Data Availability

All data produced in the present study are available upon reasonable request to the authors.

## Acknowledgements

In memory of Prof. Aart Schene, the authors are deeply grateful for his contribution in setting up the MIND-SET Study and help in formulating the research question. This work was carried out on the Dutch national e-infrastructure with the support of SURF Cooperative (Grant no. EINF1824). YS is funded by the junior researcher PhD grant from Donders Center for Medical Neuroscience at Radboudumc. ES is funded by a NARSAD Young Investigator from the Brain and Behavior Research Foundation (Grant no. 25034), and a Hypatia Tenure Track Grant (Radboudumc). NRM, BF, and JB have received funding from the European Community’s Horizon 2020 research and innovation programme under grant agreement no. 847879 (PRIME). DD was supported by the Novo Nordisk Foundation (NNF20OC0065561), the Lundbeck Foundation (R344-2020-1060). The iPSYCH team was supported by grants from the Lundbeck Foundation (R102-A9118, R155-2014-1724, and R248-2017-2003), the EU H2020 Program (Grant No. 667302, “CoCA”), NIH/NIMH (1U01MH109514-01 and 1R01MH124851-01 to ADB) and the Universities and University Hospitals of Aarhus and Copenhagen. Research reported in this publication was supported by the National Institute of Mental Health of the National Institutes of Health under Award Number R01MH124851. The content is solely the responsibility of the authors and does not necessarily represent the official views of the National Institutes of Health.

## References

1. GBD 2016 DALYs and HALE Collaborators. (2017). Global, regional, and national disability-adjusted life-years (DALYs) for 333 diseases and injuries and healthy life expectancy (HALE) for 195 countries and territories, 1990-2016: A systematic analysis for the Global Burden of Disease Study 2016. Lancet (London, England), 390(10100), 1260–1344. https://doi.org/10.1016/S0140-6736(17)32130-X

2. American Psychiatric Association. (2013). Diagnostic and statistical manual of mental disorders. American Psychiatric Publishing.

3. World Health Organization. (1993). ICD-10: The ICD-10 Classification of Mental and Behavioural Disorders: Diagnostic criteria for research. In ICD-10: The ICD-10 classification of mental and behavioural disorders: Diagnostic criteria for research (pp. xiii–248).

4. Zimmerman, M., Ellison, W., Young, D., Chelminski, I., & Dalrymple, K. (2015). How many different ways do patients meet the diagnostic criteria for major depressive disorder? Comprehensive Psychiatry, 56, 29–34. https://doi.org/10.1016/j.comppsych.2014.09.007

5. Alnæs, D., Kaufmann, T., van der Meer, D., Córdova-Palomera, A., Rokicki, J., Moberget, T., Bettella, F., Agartz, I., Barch, D. M., Bertolino, A., Brandt, C. L., Cervenka, S., Djurovic, S., Doan, N. T., Eisenacher, S., Fatouros-Bergman, H., Flyckt, L., Di Giorgio, A., Haatveit, B., … Westlye, L. T. (2019). Brain Heterogeneity in Schizophrenia and Its Association With Polygenic Risk. JAMA Psychiatry, 76(7), 739–748. https://doi.org/10.1001/jamapsychiatry.2019.0257

6. Wolfers, T., Doan, N. T., Kaufmann, T., Alnæs, D., Moberget, T., Agartz, I., Buitelaar, J. K., Ueland, T., Melle, I., Franke, B., Andreassen, O. A., Beckmann, C. F., Westlye, L. T., & Marquand, A. F. (2018). Mapping the Heterogeneous Phenotype of Schizophrenia and Bipolar Disorder Using Normative Models. JAMA Psychiatry, 75(11), 1146–1155. https://doi.org/10.1001/jamapsychiatry.2018.2467

7. Craddock, N., & Owen, M. J. (2010). The Kraepelinian dichotomy – going, going… But still not gone. The British Journal of Psychiatry, 196(2), 92–95. https://doi.org/10.1192/bjp.bp.109.073429

8. Plana-Ripoll, O., Pedersen, C. B., Holtz, Y., Benros, M. E., Dalsgaard, S., de Jonge, P., Fan, C. C., Degenhardt, L., Ganna, A., Greve, A. N., Gunn, J., Iburg, K. M., Kessing, L. V., Lee, B. K., Lim, C. C. W., Mors, O., Nordentoft, M., Prior, A., Roest, A. M., … McGrath, J. J. (2019). Exploring Comorbidity Within Mental Disorders Among a Danish National Population. JAMA Psychiatry, 76(3), 259–270. https://doi.org/10.1001/jamapsychiatry.2018.3658

9. Cuthbert, B. N., & Insel, T. R. (2013). Toward the future of psychiatric diagnosis: The seven pillars of RDoC. BMC Medicine, 11(1), 126. https://doi.org/10.1186/1741-7015-11-126

10. Kotov, R., Krueger, R. F., Watson, D., Achenbach, T. M., Althoff, R. R., Bagby, R. M., Brown, T. A., Carpenter, W. T., Caspi, A., Clark, L. A., Eaton, N. R., Forbes, M. K., Forbush, K. T., Goldberg, D., Hasin, D., Hyman, S. E., Ivanova, M. Y., Lynam, D. R., Markon, K., … Zimmerman, M. (2017). The Hierarchical Taxonomy of Psychopathology (HiTOP): A dimensional alternative to traditional nosologies. Journal of Abnormal Psychology, 126(4), 454–477. https://doi.org/10.1037/abn0000258

11. Kozak, M. J., & Cuthbert, B. N. (2016). The NIMH Research Domain Criteria Initiative: Background, Issues, and Pragmatics. Psychophysiology, 53(3), 286–297. https://doi.org/10.1111/psyp.12518

12. Wray, N. R., Ripke, S., Mattheisen, M., Trzaskowski, M., Byrne, E. M., Abdellaoui, A., Adams, M. J., Agerbo, E., Air, T. M., Andlauer, T. M. F., Bacanu, S.-A., Bækvad-Hansen, M., Beekman, A. F. T., Bigdeli, T. B., Binder, E. B., Blackwood, D. R. H., Bryois, J., Buttenschøn, H. N., Bybjerg-Grauholm, J., … Sullivan, P. F. (2018). Genome-wide association analyses identify 44 risk variants and refine the genetic architecture of major depression. Nature Genetics, 50(5), 668–681. https://doi.org/10.1038/s41588-018-0090-3

13. Purves, K. L., Coleman, J. R. I., Meier, S. M., Rayner, C., Davis, K. A. S., Cheesman, R., Bækvad-Hansen, M., Børglum, A. D., Wan Cho, S., Jürgen Deckert, J., Gaspar, H. A., Bybjerg-Grauholm, J., Hettema, J. M., Hotopf, M., Hougaard, D., Hübel, C., Kan, C., McIntosh, A. M., Mors, O., … Eley, T. C. (2020). A major role for common genetic variation in anxiety disorders. Molecular Psychiatry, 25(12), 3292–3303. https://doi.org/10.1038/s41380-019-0559-1

14. Demontis, D., Walters, G. B., Athanasiadis, G., Walters, R., Therrien, K., Farajzadeh, L., Voloudakis, G., Bendl, J., Zeng, B., Zhang, W., Grove, J., Als, T. D., Duan, J., Satterstrom, F. K., Bybjerg-Grauholm, J., Bækved-Hansen, M., Gudmundsson, O. O., Magnusson, S. H., Baldursson, G., … Børglum, A. D. (2022). Genome-wide analyses of ADHD identify 27 risk loci, refine the genetic architecture and implicate several cognitive domains (p. 2022.02.14.22270780). medRxiv. https://doi.org/10.1101/2022.02.14.22270780

15. Grove, J., Ripke, S., Als, T. D., Mattheisen, M., Walters, R. K., Won, H., Pallesen, J., Agerbo, E., Andreassen, O. A., Anney, R., Awashti, S., Belliveau, R., Bettella, F., Buxbaum, J. D., Bybjerg-Grauholm, J., Bækvad-Hansen, M., Cerrato, F., Chambert, K., Christensen, J. H., … Børglum, A. D. (2019). Identification of common genetic risk variants for autism spectrum disorder. Nature Genetics, 51(3), 431–444. https://doi.org/10.1038/s41588-019-0344-8

16. Mullins, N., Forstner, A. J., O’Connell, K. S., Coombes, B., Coleman, J. R. I., Qiao, Z., Als, T. D., Bigdeli, T. B., Børte, S., Bryois, J., Charney, A. W., Drange, O. K., Gandal, M. J., Hagenaars, S. P., Ikeda, M., Kamitaki, N., Kim, M., Krebs, K., Panagiotaropoulou, G., … Andreassen, O. (2021). Genome-wide association study of over 40,000 bipolar disorder cases provides new insights into the underlying biology (p. 2020.09.17.20187054). medRxiv. https://doi.org/10.1101/2020.09.17.20187054

17. Pardiñas, A. F., Holmans, P., Pocklington, A. J., Escott-Price, V., Ripke, S., Carrera, N., Legge, S. E., Bishop, S., Cameron, D., Hamshere, M. L., Han, J., Hubbard, L., Lynham, A., Mantripragada, K., Rees, E., MacCabe, J. H., McCarroll, S. A., Baune, B. T., Breen, G., … Walters, J. T. R. (2018). Common schizophrenia alleles are enriched in mutation-intolerant genes and in regions under strong background selection. Nature Genetics, 50(3), 381–389. https://doi.org/10.1038/s41588-018-0059-2

18. The Brainstorm Consortium, Anttila, V., Bulik-Sullivan, B., Finucane, H. K., Walters, R. K., Bras, J., Duncan, L., Escott-Price, V., Falcone, G. J., Gormley, P., Malik, R., Patsopoulos, N. A., Ripke, S., Wei, Z., Yu, D., Lee, P. H., Turley, P., Grenier-Boley, B., Chouraki, V., … Neale, B. M. (2018). Analysis of shared heritability in common disorders of the brain. Science, 360(6395), eaap8757. https://doi.org/10.1126/science.aap8757

19. Grotzinger, A. D., Mallard, T. T., Akingbuwa, W. A., Ip, H. F., Adams, M. J., Lewis, C. M., McIntosh, A. M., Grove, J., Dalsgaard, S., Lesch, K.-P., Strom, N., Meier, S. M., Mattheisen, M., Børglum, A. D., Mors, O., Breen, G., Lee, P. H., Kendler, K. S., Smoller, J. W., … Nivard, M. G. (2022). Genetic architecture of 11 major psychiatric disorders at biobehavioral, functional genomic and molecular genetic levels of analysis. Nature Genetics, 54(5), 548–559. https://doi.org/10.1038/s41588-022-01057-4

20. Lee, P. H., Anttila, V., Won, H., Feng, Y.-C. A., Rosenthal, J., Zhu, Z., Tucker-Drob, E. M., Nivard, M. G., Grotzinger, A. D., Posthuma, D., Wang, M. M.-J., Yu, D., Stahl, E. A., Walters, R. K., Anney, R. J. L., Duncan, L. E., Ge, T., Adolfsson, R., Banaschewski, T., … Smoller, J. W. (2019). Genomic Relationships, Novel Loci, and Pleiotropic Mechanisms across Eight Psychiatric Disorders. Cell, 179(7), 1469-1482.e11. https://doi.org/10.1016/j.cell.2019.11.020

21. Khera, A. V., Chaffin, M., Aragam, K. G., Haas, M. E., Roselli, C., Choi, S. H., Natarajan, P., Lander, E. S., Lubitz, S. A., Ellinor, P. T., & Kathiresan, S. (2018). Genome-wide polygenic scores for common diseases identify individuals with risk equivalent to monogenic mutations. Nature Genetics, 50(9), 1219–1224. https://doi.org/10.1038/s41588-018-0183-z

22. Luykx, J. J., Loef, D., Lin, B., van Diermen, L., Nuninga, J. O., van Exel, E., Oudega, M. L., Rhebergen, D., Schouws, S. N. T. M., van Eijndhoven, P., Verwijk, E., Schrijvers, D., Birkenhager, T. K., Ryan, K. M., Arts, B., van Bronswijk, S. C., Kenis, G., Schurgers, G., Baune, B. T., … Rutten, B. P. F. (2022). Interrogating Associations Between Polygenic Liabilities and Electroconvulsive Therapy Effectiveness. Biological Psychiatry, 91(6), 531–539. https://doi.org/10.1016/j.biopsych.2021.10.013

23. Cross-Disorder Group of the Psychiatric Genomics Consortium. (2013). Identification of risk loci with shared effects on five major psychiatric disorders: A genome-wide analysis. Lancet (London, England), 381(9875), 1371–1379. https://doi.org/10.1016/S0140-6736(12)62129-1

24. Bralten, J., Mota, N. R., Klemann, C. J. H. M., De Witte, W., Laing, E., Collier, D. A., de Kluiver, H., Bauduin, S. E. E. C., Arango, C., Ayuso-Mateos, J. L., Fabbri, C., Kas, M. J., van der Wee, N., Penninx, B. W. J. H., Serretti, A., Franke, B., & Poelmans, G. (2021). Genetic underpinnings of sociability in the general population. Neuropsychopharmacology: Official Publication of the American College of Neuropsychopharmacology, 46(9), 1627–1634. https://doi.org/10.1038/s41386-021-01044-z

25. Howard, D. M., Adams, M. J., Clarke, T.-K., Hafferty, J. D., Gibson, J., Shirali, M., Coleman, J. R. I., Hagenaars, S. P., Ward, J., Wigmore, E. M., Alloza, C., Shen, X., Barbu, M. C., Xu, E. Y., Whalley, H. C., Marioni, R. E., Porteous, D. J., Davies, G., Deary, I. J., … McIntosh, A. M. (2019). Genome-wide meta-analysis of depression identifies 102 independent variants and highlights the importance of the prefrontal brain regions. Nature Neuroscience, 22(3), 343–352. https://doi.org/10.1038/s41593-018-0326-7

26. Eijndhoven, P. van, Collard, R., Vrijsen, J., Geurts, D. E. M., Vasquez, A. A., Schellekens, A., Munckhof, E. van den, Brolsma, S., Duyser, F., Bergman, A., Oort, J. van, Tendolkar, I., & Schene, A. (2022). Measuring Integrated Novel Dimensions in Neurodevelopmental and Stress-Related Mental Disorders (MIND-SET): Protocol for a Cross-sectional Comorbidity Study From a Research Domain Criteria Perspective. JMIRx Med, 3(1), e31269. https://doi.org/10.2196/31269

27. First, M., Gibbon, M., Spitzer, R., Williams, J., & Benjamin, L. (1997). Structured clinical interview for DSM-IV-TR axis I disorders, research version, patient edition. (SCID-I/P). New York: Biometrics Research, New York State Psychiatric Institute.

28. Vuijk, R. (2016). Nederlands interview ten behoeve van diagnostiek autismespectrumstoornis bij volwassenen (NIDA). Rotterdam: Sarr Expertisecentrum Autisme/Dare to Design.

29. Kooij, J. J. S., & Francken, M. H. (2010). Diagnostic Interview for ADHD in adults (DIVA). DIVA Foundation, The Netherlands.

30. Schippers, G. M., Broekman, T. G., Buchholz, A., Koeter, M. W. J., & van den Brink, W. (2010). Measurements in the Addictions for Triage and Evaluation (MATE): An instrument based on the World Health Organization family of international classifications. Addiction (Abingdon, England), 105(5), 862–871. https://doi.org/10.1111/j.1360-0443.2009.02889.x

31. World Health Organization Disability Assessment Schedule 2.0. (2017). In A. Wenzel, The SAGE Encyclopedia of Abnormal and Clinical Psychology. SAGE Publications, Inc. https://doi.org/10.4135/9781483365817.n1493

32. de Jong, K., Nugter, M. A., Polak, M. G., Wagenborg, J. E. A., Spinhoven, P., & Heiser, W. J. (2007). The Outcome Questionnaire (OQ-45) in a Dutch population: A cross-cultural validation. Clinical Psychology & Psychotherapy, 14(4), 288–301. https://doi.org/10.1002/cpp.529

33. Mulders, P., van Eijndhoven, P., van Oort, J., Oldehinkel, M., Duyser, F., Kist, J., Collard, R., Vrijsen, J., Haak, K., Beckmann, C., Tendolkar, I., & Marquand, A. (n.d.). Striatal connectopic maps link to functional domains across psychiatric disorders.

34. Horn, J. L. (1965). A rationale and test for the number of factors in factor analysis. Psychometrika, 30(2), 179–185. https://doi.org/10.1007/BF02289447

35. Howard, D. M., Adams, M. J., Shirali, M., Clarke, T.-K., Marioni, R. E., Davies, G., Coleman, J. R. I., Alloza, C., Shen, X., Barbu, M. C., Wigmore, E. M., Gibson, J., 23andMe Research Team, Hagenaars, S. P., Lewis, C. M., Ward, J., Smith, D. J., Sullivan, P. F., Haley, C. S., … McIntosh, A. M. (2018). Genome-wide association study of depression phenotypes in UK Biobank identifies variants in excitatory synaptic pathways. Nature Communications, 9(1), 1470. https://doi.org/10.1038/s41467-018-03819-3

36. Lam, M., Awasthi, S., Watson, H. J., Goldstein, J., Panagiotaropoulou, G., Trubetskoy, V., Karlsson, R., Frei, O., Fan, C.-C., De Witte, W., Mota, N. R., Mullins, N., Brügger, K., Lee, S. H., Wray, N. R., Skarabis, N., Huang, H., Neale, B., Daly, M. J., … Ripke, S. (2020). RICOPILI: Rapid Imputation for COnsortias PIpeLIne. Bioinformatics, 36(3), 930–933. https://doi.org/10.1093/bioinformatics/btz633

37. Kalia, S. S., Adelman, K., Bale, S. J., Chung, W. K., Eng, C., Evans, J. P., Herman, G. E., Hufnagel, S. B., Klein, T. E., Korf, B. R., & others. (2017). Recommendations for reporting of secondary findings in clinical exome and genome sequencing, 2016 update (ACMG SF v2. 0): A policy statement of the American College of Medical Genetics and Genomics. Genetics in Medicine, 19(2), 249–255.

38. Fairley, S., Lowy-Gallego, E., Perry, E., & Flicek, P. (2020). The International Genome Sample Resource (IGSR) collection of open human genomic variation resources. Nucleic Acids Research, 48(D1), D941–D947. https://doi.org/10.1093/nar/gkz836

39. Das, S., Forer, L., Schönherr, S., Sidore, C., Locke, A. E., Kwong, A., Vrieze, S. I., Chew, E. Y., Levy, S., McGue, M., Schlessinger, D., Stambolian, D., Loh, P.-R., Iacono, W. G., Swaroop, A., Scott, L. J., Cucca, F., Kronenberg, F., Boehnke, M., … Fuchsberger, C. (2016). Next-generation genotype imputation service and methods. Nature Genetics, 48(10), 1284–1287. https://doi.org/10.1038/ng.3656

40. Choi, S. W., & O’Reilly, P. F. (2019). PRSice-2: Polygenic Risk Score software for biobank-scale data. GigaScience, 8(7), giz082. https://doi.org/10.1093/gigascience/giz082

41. Cai, N., Revez, J. A., Adams, M. J., Andlauer, T. F. M., Breen, G., Byrne, E. M., Clarke, T.-K., Forstner, A. J., Grabe, H. J., Hamilton, S. P., Levinson, D. F., Lewis, C. M., Lewis, G., Martin, N. G., Milaneschi, Y., Mors, O., Müller-Myhsok, B., Penninx, B. W. J. H., Perlis, R. H., … Flint, J. (2020). Minimal phenotyping yields genome-wide association signals of low specificity for major depression. Nature Genetics, 52(4), 437–447. https://doi.org/10.1038/s41588-020-0594-5

42. Mitchell, B. L., Thorp, J. G., Wu, Y., Campos, A. I., Nyholt, D. R., Gordon, S. D., Whiteman, D. C., Olsen, C. M., Hickie, I. B., Martin, N. G., Medland, S. E., Wray, N. R., & Byrne, E. M. (2021). Polygenic Risk Scores Derived From Varying Definitions of Depression and Risk of Depression. JAMA Psychiatry, 78(10), 1152–1160. https://doi.org/10.1001/jamapsychiatry.2021.1988

43. McCoy, T. H., Yu, S., Hart, K. L., Castro, V. M., Brown, H. E., Rosenquist, J. N., Doyle, A. E., Vuijk, P. J., Cai, T., & Perlis, R. H. (2018). High Throughput Phenotyping for Dimensional Psychopathology in Electronic Health Records. Biological Psychiatry, 83(12), 997–1004. https://doi.org/10.1016/j.biopsych.2018.01.011

44. de la Fuente, J., Davies, G., Grotzinger, A. D., Tucker-Drob, E. M., & Deary, I. J. (2021). A general dimension of genetic sharing across diverse cognitive traits inferred from molecular data. Nature Human Behaviour, 5(1), 49–58. https://doi.org/10.1038/s41562-020-00936-2

45. Genetics of Personality Consortium. (2015). Meta-analysis of Genome-wide Association Studies for Neuroticism, and the Polygenic Association With Major Depressive Disorder. JAMA Psychiatry, 72(7), 642–650. https://doi.org/10.1001/jamapsychiatry.2015.0554

46. Klein Hofmeijer-Sevink, M., Batelaan, N. M., van Megen, H. J. G. M., Penninx, B. W., Cath, D. C., van den Hout, M. A., & van Balkom, A. J. L. M. (2012). Clinical relevance of comorbidity in anxiety disorders: A report from the Netherlands Study of Depression and Anxiety (NESDA). Journal of Affective Disorders, 137(1), 106–112. https://doi.org/10.1016/j.jad.2011.12.008

47. Overbeek, T., Schruers, K., & Griez, E. (2002). Comorbidity of Obsessive-Compulsive Disorder and Depression: Prevalence, Symptom Severity, and Treatment Effect. The Journal of Clinical Psychiatry, 63(12), 3395.

