## Supplementary Tables and Figures for "Multi-polygenic scores in psychiatry: from disorder-specific to transdiagnostic perspectives"

Table S1. Overview of diagnosis and grouping in the MIND-SET cohort

| **Disorder group** | Diagnosis | Sample size | | |
| --- | --- | --- | --- | --- |
| **Mood Disorders** | Past unipolar depression | 453 | 360 | 276 |
|  | Current dysthymia |  |  | 54 |
|  | Current depressive episode |  |  | 195 |
|  | Current (hypo)manic episode |  |  | 4 |
|  | Bipolar disorder |  |  | 21 |
| **Anxiety Disorders** | Panic disorder |  | 147 | 33 |
|  | Agoraphobia |  |  | 4 |
|  | Social phobia |  |  | 44 |
|  | Specific phobia |  |  | 11 |
|  | Obsessive-compulsive disorder |  |  | 16 |
|  | Post-traumatic stress disorder |  |  | 35 |
|  | Generalized anxiety disorder |  |  | 28 |
|  | Anxiety not otherwise specified |  |  | 18 |
| **ADHD** | Attention-deficit/hyperactivity disorder |  | 171 | |
| **ASD** | Autism spectrum disorder |  | 121 | |
| **SUD** | Substance use disorders |  | 121 | |

**Table S2. Questionnaire subscales included in factor analysis.**

| Symptoms/ Traits | Questionnaires | Subscales | |
| --- | --- | --- | --- |
| Depressive symptoms | Inventory of Depressive Symptomatology -Self Rating (IDS-SR) [1] | No subscales | |
| Anxiety sensitivity | Anxiety Sensitivity Index (ASI) [2] | No subscales | |
| ADHD symptom severity | Conners’ Adult ADHD Rating Scale (CAARS) [3] | Inattention/memory problems  Hyperactivity/restlessness  Impulsivity/emotional liability  Problems with self-concept | |
| Autistic traits | Autism Spectrum Quotient-50 (AQ-50) [4] | Social insight and behavior  Switching attention/difficulty with changes  Communication  Fantasy/Imagination  Detail-oriented | |
| Personality traits | Personality Inventory for DSM-5-Short Form (PID-5-SF) [5] | Negative affect  Detachment  Antagonism  Disinhibition  Psychoticism | |
| Psychological constructs:   - Alexithymia - Behavioral regulation - Repetitive thoughts | Toronto Alexithymia Scale-20 (TAS-20) [6] | Difficulty describing feelings  Difficulty identifying feelings  Externally-oriented thinking | |
|  | Behavior Rating Inventory Executive Function – Adult (BRIEF-A) [7] | Inhibit  Shift  Emotional Control  Self-Monitor  Initiate | Working Memory  Plan/Organise  Organisation of Materials  Task Monitor |
|  | Perseverative Thinking Questionnaire (PTQ) [8] | Repetitiveness  Intrusiveness  difficulties to disengage | |

**Table S3. Overview of the sources of PRS base GWAS**

|  | **Phenotype** | **Abbreviation** | **Cohort** | **#Cases** | **#Controls** |
| --- | --- | --- | --- | --- | --- |
| 1 | Major depressive disorder [9] | MDD | 29 PGC MDD2 cohorts, deCODE, Generation Scotland, GERA, and iPSYCH cohorts | 45,396 | 97,250 |
| 2 | Anxiety disorders [10] | ANX | UKBB, iPSYCH, ANGST | 25,453 | 58,113 |
| 3 | Attention-deficit/hyperactivity disorder [11] | ADHD | iPSYCH + 10 PGC cohorts + deCODE | 38,691 | 186,843 |
| 4 | Autism spectrum disorder [12] | ASD | 5 PGC trio samples + iPSYCH | 18,382 | 27,969 |
| 5 | Bipolar disorder [13] | BP | PGC 57 cohorts | 41,917 | 371,549 |
| 6 | Schizophrenia [14] | SCZ | CLOZUK+PGC2 | 40,675 | 64,643 |
| 7 | Broadly defined depression [15] | DEP | 33 PGC cohorts + broad depression phenotype in UKB | 170,756 | 329,443 |
| 8 | Eight psychiatric disorders: MDD, ADHD, ASD, BP, SCZ, anorexia nervosa, obsessive-compulsive disorder, Tourette syndrome [16] | Cross-disorder | Aggregated from eight GWASs | 162,151 | 276,846 |

Table S4. Comparing generalized linear regression models with multi-PRSs and with single predictor of DEP-PRS

Model 1: Outcome ~ MDD.PRS + ANX.PRS + ADHD.PRS + ASD.PRS + BP.PRS + SCZ.PRS + DEP.PRS + Cross-disorder.PRS + C1 + C2 + C3 + C4 + Age + Sex

Model 2: Outcome ~ DEP.PRS + C1 + C2 + C3 + C4 + Age + Sex

| **Outcome** | **Df** | **Deviance** | **Pr(>Chi)** |
| --- | --- | --- | --- |
| **Mood disorders** | 7 | 10.922 | 0.142 |
| **Anxiety disorders** | 7 | 7.869 | 0.344 |
| **ADHD** | 7 | 4.768 | 0.688 |
| **ASD** | 7 | 3.844 | 0.798 |
| **SUD** | 7 | 10.969 | 0.140 |

Table S5. Multiple linear regression of individuals’ factor loadings on OQ-45 and WHODAS 2.0 scales

Linear hypotheses: OQ ~ F1 + F2 + F3 + F4

| **OQ** | **Estimate** | **SE** | **t value** | **Pr(>\|t\|)** |
| --- | --- | --- | --- | --- |
| **(Intercept)** | 69.5002 | 0.6563 | 105.896 | < 2e-16 |
| **F1** | 21.6729 | 0.8041 | 26.952 | < 2e-16 |
| **F2** | 4.0519 | 0.715 | 5.667 | 2.86e-08 |
| **F3** | 3.8304 | 0.7632 | 5.019 | 7.98e-07 |
| **F4** | 2.6194 | 0.7315 | 3.581 | 0.000386 |

Linear hypotheses: WHODAS ~ F1 + F2 + F3 + F4

| **WHODAS** | **Estimate** | **SE** | **t value** | **Pr(>\|t\|)** |
| --- | --- | --- | --- | --- |
| **(Intercept)** | 39.2391 | 0.7713 | 50.875 | < 2e-16 |
| **F1** | 15.1811 | 0.9462 | 16.044 | < 2e-16 |
| **F2** | 3.7998 | 0.8412 | 4.517 | 8.35e-06 |
| **F3** | 6.0577 | 0.8962 | 6.759 | 5.16e-11 |
| **F4** | 1.0329 | 0.8586 | 1.203 | 0.23 |

Table S6. T-test statistics for factor loadings of patients versus controls

|  | **t value** | **Pr(>\|t\|)** | 95% CI |
| --- | --- | --- | --- |
| **F1** | -16.25 | <2.2e-16 | [-1.97, -1.55] |
| **F2** | -9.46 | <2.2e-16 | [-1.45, -0.95] |
| **F3** | -7.34 | 1.32e-12 | [-1.23, -0.71] |
| **F4** | -9.55 | <2.2e-16 | [-1.46, -0.96] |

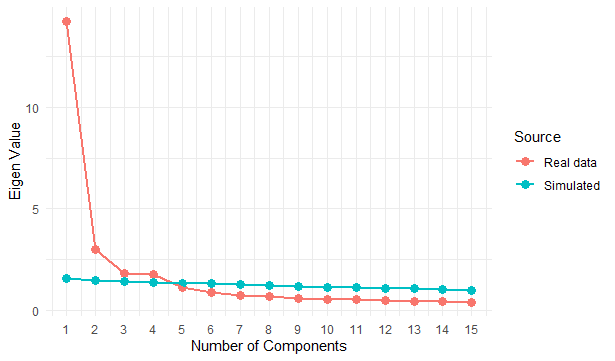

**Figure S1. Scree plot from parallel analysis.** Simulated eigenvalues were calculated from 500 randomly generated correlation matrices, which were then compared with eigenvalues extracted from the real data [17]. The 95th percentile of eigenvalues were used for this comparison, and the four factors larger than the simulated eigenvalues were retained. This analysis was performed using an R-based engine [18].

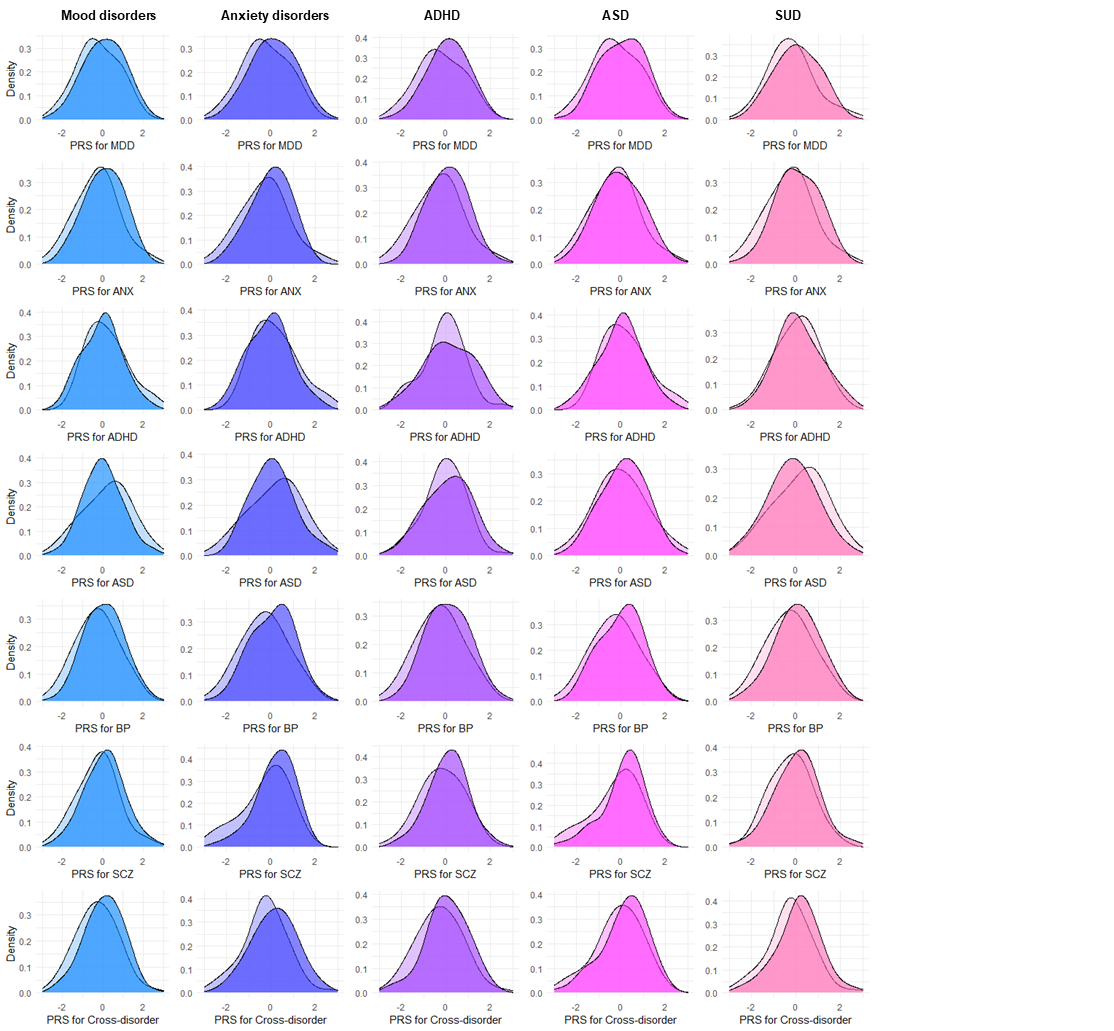
**Figure S2**. **PRS distributions for unaffected (light color) and affected individuals (dark color).** Each row shows the liability computed per base GWAS (denoted in the x-axis label). Each column shows the PRS distributions of individuals with or without the diagnosis per disorder (denoted as the column header).

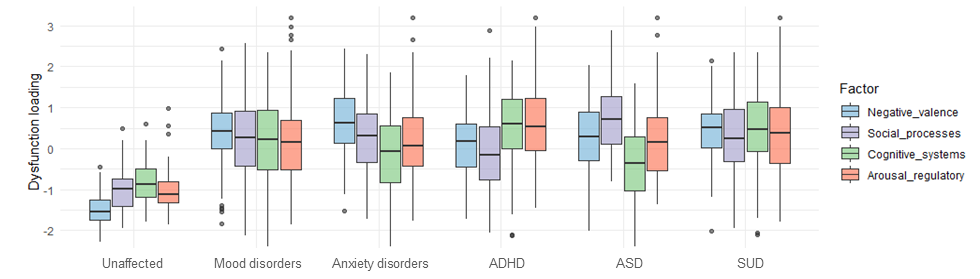
**Figure S3. Factor loadings per diagnostic group.** The median per factor per group is shown in each box plot; the lower and upper hinges correspond to the 1^st^ and 3^rd^ quartiles; the whiskers extend from the hinge to the largest/smallest values no further than 1.5 times the inter-quartile range. Outliers are plotted individually.

**References**

1. Rush, A. J., Gullion, C. M., Basco, M. R., Jarrett, R. B., & Trivedi, M. H. (1996). The Inventory of Depressive Symptomatology (IDS): Psychometric properties. *Psychological Medicine*, *26*(3), 477–486. <https://doi.org/10.1017/S0033291700035558>
2. Reiss, S., Peterson, R. A., Gursky, D. M., & McNally, R. J. (1986). Anxiety sensitivity, anxiety frequency and the prediction of fearfulness. *Behaviour Research and Therapy*, *24*(1), 1–8. <https://doi.org/10.1016/0005-7967(86)90143-9>
3. Conners, C. K., Erhardt, D., & Sparrow, E. P. (1999). *Conners' adult ADHD rating scales (CAARS): technical manual*. North Tonawanda, NY: Multi-Health Systems.
4. Baron-Cohen, S., Wheelwright, S., Skinner, R., Martin, J., & Clubley, E. (2001). The Autism-Spectrum Quotient (AQ): Evidence from Asperger Syndrome/High-Functioning Autism, Malesand Females, Scientists and Mathematicians. *Journal of Autism and Developmental Disorders*, *31*(1), 5–17. <https://doi.org/10.1023/A:1005653411471>
5. Díaz-Batanero, C., Ramírez-López, J., Domínguez-Salas, S., Fernández-Calderón, F., & Lozano, Ó. M. (2019). Personality Inventory for DSM-5–Short Form (PID-5-SF): Reliability, Factorial Structure, and Relationship With Functional Impairment in Dual Diagnosis Patients. *Assessment*, *26*(5), 853–866. <https://doi.org/10.1177/1073191117739980>
6. Leising, D., Grande, T., & Faber, R. (2009). The Toronto Alexithymia Scale (TAS-20): A measure of general psychological distress. *Journal of Research in Personality*, *43*(4), 707–710. <https://doi.org/10.1016/j.jrp.2009.03.009>
7. Roth, R. M., & Gioia, G. A. (2005). *Behavior rating inventory of executive function--adult version*. Lutz, FL: Psychological Assessment Resources.
8. Ehring, T., Zetsche, U., Weidacker, K., Wahl, K., Schönfeld, S., & Ehlers, A. (2011). The Perseverative Thinking Questionnaire (PTQ): Validation of a content-independent measure of repetitive negative thinking. *Journal of Behavior Therapy and Experimental Psychiatry*, *42*(2), 225–232. <https://doi.org/10.1016/j.jbtep.2010.12.003>
9. Wray, N. R., Ripke, S., Mattheisen, M., Trzaskowski, M., Byrne, E. M., Abdellaoui, A., Adams, M. J., Agerbo, E., Air, T. M., Andlauer, T. M. F., Bacanu, S.-A., Bækvad-Hansen, M., Beekman, A. F. T., Bigdeli, T. B., Binder, E. B., Blackwood, D. R. H., Bryois, J., Buttenschøn, H. N., Bybjerg-Grauholm, J., … Sullivan, P. F. (2018). Genome-wide association analyses identify 44 risk variants and refine the genetic architecture of major depression. *Nature Genetics*, *50*(5), 668–681. <https://doi.org/10.1038/s41588-018-0090-3>
10. Purves, K. L., Coleman, J. R. I., Meier, S. M., Rayner, C., Davis, K. A. S., Cheesman, R., Bækvad-Hansen, M., Børglum, A. D., Wan Cho, S., Jürgen Deckert, J., Gaspar, H. A., Bybjerg-Grauholm, J., Hettema, J. M., Hotopf, M., Hougaard, D., Hübel, C., Kan, C., McIntosh, A. M., Mors, O., … Eley, T. C. (2020). A major role for common genetic variation in anxiety disorders. *Molecular Psychiatry*, *25*(12), 3292–3303. <https://doi.org/10.1038/s41380-019-0559-1>
11. Demontis, D., Walters, B., Athanasiadis, G., Walters, R., Therrien, K., Farajzadeh, L., ... & Boerglum, A. D. (2022). Genome-wide analyses of ADHD identify 27 risk loci, refine the genetic architecture and implicate several cognitive domains. *medRxiv*.
12. Grove, J., Ripke, S., Als, T. D., Mattheisen, M., Walters, R. K., Won, H., Pallesen, J., Agerbo, E., Andreassen, O. A., Anney, R., Awashti, S., Belliveau, R., Bettella, F., Buxbaum, J. D., Bybjerg-Grauholm, J., Bækvad-Hansen, M., Cerrato, F., Chambert, K., Christensen, J. H., … Børglum, A. D. (2019). Identification of common genetic risk variants for autism spectrum disorder. *Nature Genetics*, *51*(3), 431–444. <https://doi.org/10.1038/s41588-019-0344-8>
13. Mullins, N., Forstner, A. J., O’Connell, K. S., Coombes, B., Coleman, J. R. I., Qiao, Z., Als, T. D., Bigdeli, T. B., Børte, S., Bryois, J., Charney, A. W., Drange, O. K., Gandal, M. J., Hagenaars, S. P., Ikeda, M., Kamitaki, N., Kim, M., Krebs, K., Panagiotaropoulou, G., … Andreassen, O. A. (2021). Genome-wide association study of more than 40,000 bipolar disorder cases provides new insights into the underlying biology. *Nature Genetics*, *53*(6), 817–829. <https://doi.org/10.1038/s41588-021-00857-4>
14. Pardiñas, A. F., Holmans, P., Pocklington, A. J., Escott-Price, V., Ripke, S., Carrera, N., Legge, S. E., Bishop, S., Cameron, D., Hamshere, M. L., Han, J., Hubbard, L., Lynham, A., Mantripragada, K., Rees, E., MacCabe, J. H., McCarroll, S. A., Baune, B. T., Breen, G., … Walters, J. T. R. (2018). Common schizophrenia alleles are enriched in mutation-intolerant genes and in regions under strong background selection. *Nature Genetics*, *50*(3), 381–389. <https://doi.org/10.1038/s41588-018-0059-2>
15. Howard, D. M., Adams, M. J., Clarke, T.-K., Hafferty, J. D., Gibson, J., Shirali, M., Coleman, J. R. I., Hagenaars, S. P., Ward, J., Wigmore, E. M., Alloza, C., Shen, X., Barbu, M. C., Xu, E. Y., Whalley, H. C., Marioni, R. E., Porteous, D. J., Davies, G., Deary, I. J., … McIntosh, A. M. (2019). Genome-wide meta-analysis of depression identifies 102 independent variants and highlights the importance of the prefrontal brain regions. *Nature Neuroscience*, *22*(3), 343–352. <https://doi.org/10.1038/s41593-018-0326-7>
16. Lee, P. H., Anttila, V., Won, H., Feng, Y.-C. A., Rosenthal, J., Zhu, Z., Tucker-Drob, E. M., Nivard, M. G., Grotzinger, A. D., Posthuma, D., Wang, M. M.-J., Yu, D., Stahl, E. A., Walters, R. K., Anney, R. J. L., Duncan, L. E., Ge, T., Adolfsson, R., Banaschewski, T., … Smoller, J. W. (2019). Genomic Relationships, Novel Loci, and Pleiotropic Mechanisms across Eight Psychiatric Disorders. *Cell*, *179*(7), 1469-1482.e11. <https://doi.org/10.1016/j.cell.2019.11.020>
17. Horn, J. L. (1965). A rationale and test for the number of factors in factor analysis. *Psychometrika, 30*(2), 179–185. <https://doi.org/10.1007/BF02289447>
18. Patil Vivek H, Surendra N. Singh, Sanjay Mishra, and D. Todd Donavan (2017). Parallel Analysis Engine to Aid in Determining Number of Factors to Retain using R [Computer software], available from <https://analytics.gonzaga.edu/parallelengine/>.
